# Patient-specific analysis of bicuspid aortic valve hemodynamics using a fully coupled fluid-structure interaction (FSI) model

**DOI:** 10.1101/2021.10.24.21265224

**Authors:** Tongran Qin, Andrés Caballero, Wenbin Mao, Brian Barrett, Norihiko Kamioka, Stamatios Lerakis, Wei Sun

**Author notes:** For correspondence Wei Sun, PhD, The Wallace H. Coulter Department of Biomedical Engineering, Georgia Institute of Technology and Emory University, Technology Enterprise Park, Room 206, 387 Technology Circle, Atlanta, GA 30313-2412.

## Abstract

Bicuspid aortic valve (BAV), the most common congenital heart disease, is prone to develop significant valvular dysfunction and aortic wall abnormalities. Growing evidence has suggested that abnormal BAV hemodynamics could contribute to the disease progression. In order to investigate the BAV hemodynamic, we performed 3D patient-specific fluid-structure interaction (FSI) simulations of BAV with fully coupled flow dynamics and valve motions throughout the cardiac cycle. The results showed that the flow during systole can be characterized by a systolic jet and two counter-rotating recirculation vortices. At peak systole, the jet was usually eccentric, with asymmetric recirculation vortices, and helical flow motion in the ascending aorta. The flow structure at peak systole was quantified using the vorticity, flow reversal ratio and helicity index at four locations from the aortic root to the ascending aorta. The systolic jet was evaluated using the metrics including the peak velocity, normalized flow displacement, and jet angle. It was found that both the peak velocity and normalized flow displacement (rather than jet angle) of the systolic jet showed a strong correlation with the vorticity and helicity index of the flow in the ascending aorta, which suggests that these two metrics can be used for noninvasive evaluation of abnormal flow patterns in BAV patients.

## 1. INTRODUCTION

Bicuspid aortic valve (BAV) is considered the most common congenital heart disease, occurring in 0.5–2% of the population (1-3), where the aortic valve (AV) has only two rather than the normal three leaflets. Compared with a tricuspid AV (TAV), BAV patients are more prone to harbor not only significant valvular dysfunction such as aortic stenosis (AS) and aortic insufficiency (AI), but also aortic wall abnormalities such as the ascending aortic dilation (aortopathy) and aneurysms (2). While the exact cause of diseases associated with BAV remains unclear, as both genetic factors and biomechanical factors can play a role, growing evidence has shown that the abnormal hemodynamics of BAV could contribute to disease progression (4-7). For example, recent studies have found that BAV aortopathy is different from genetic aortopathy found in Marfan patients (8). Both *in vivo* data analysis (5, 9) and numerical modeling of BAV (10-13) have reported a more eccentric systolic jet compared to TAV. This asymmetry in the flow was found to associate with an elevated regional wall shear stress and the development of BAV aortopathy (6). In addition, while normal helical flow could facilitate blood flow transport and protect against atherosclerosis (14, 15), the more intense helical flows identified in BAV patients were linked to the development of aortic aneurysms (16-18).

Computational modeling has provided an engineering approach for detailed interrogation of hemodynamic factors related to BAV. However, most of the computational BAV models have been significantly limited by their inability to simultaneously solve the soft tissue mechanics and the fluid dynamics, as many of them only considered the blood flow without the consideration of the valve deformation (11-13, 19). While fluid-structure interaction (FSI) models were developed in recently studies, many of them were still limited to idealized leaflet geometries (20-22) or simplified 2D setup (23, 24). With these challenges in mind, the objectives of this study are to: 1) develop and validate comprehensive FSI models of patient-specific BAVs, 2) analyze the BAV hemodynamics in the aortic root and the ascending aorta over the cardiac cycle, and 3) quantify the flow patterns at peak systole, and investigate their relation with multiple jet metrics. Following our previous structural analysis of patient specific BAVs (25), we created five patient-specific FSI models of BAV patients from multi-slice computed tomography (MSCT) images, which included the BAV (with raphe and calcifications), the aortic root, the ascending aorta, proximal left ventricle (LV) and the surrounding myocardium. The FSI simulations were performed to simulate the valve movement and the blood flow over the entire cardiac cycle. Flow patterns were quantified using the magnitude of vorticity, flow reversal ratio and helicity index at different distal locations from the sinus level to the ascending aorta. The systolic jet was evaluated with multiple jet metrics including the peak velocity, normalized flow displacement and jet angle. The correlations between the parameters that quantified the flow patterns in the ascending aorta and the multiple jet metrics were investigated.

## 2. METHODS

### 2.1 Patient clinical data

Clinical data from five BAV patients between the age of 57 and 82 were collected retrospectively from Emory University Hospital (Atlanta, GA) with institutional Review Broad (IRB) approval (Table 1). Following the BAV classification system by Sievers and Schmidtke (26) (Figure 2a), two patients had type 0 BAV (without raphe) and three patients had type 1 BAV (with one raphe). All BAV patients had severe AS with moderate to severe calcifications. The MSCT images were acquired using a SIEMENS SOMATOM Definition Flash CT scanner with an in-plane spatial resolution between 0.62 × 0.62 - 0.93 × 0.93 mm, and a slice thickness of 1.0 mm.

**Table 1.** Patient information: BAV type, peak transvalvular diastolic and systolic pressures. Type 0 and type 1 refers to a BAV with no raphe, and one raphe, respectively. L, R, and N refers to the left, right, and non-coronary leaflets, respectively. A-P refers to a BAV where the orientation of the leaflet free edge is anterior-posterior, LAT refers to a BAV where the orientation of the leaflet free edge is lateral. L-R refers to a BAV where the left and right coronary leaflets are fused, while R-N refers to a BAV where the right and non-coronary leaflets are fused. A schematic of different types of BAV can be found in Figure 2a.

| Patient ID | Type of BAV | Transvalvular diastolic pressure gradient (mmHg) | Transvalvular systolic pressure gradient (mmHg) |
| --- | --- | --- | --- |
| A | 0 A-P | 57 | 90 |
| B | 0 LAT | 60 | 106 |
| C | 1 R-N | 54 | 65 |
| D | 1 L-R | 77 | 95 |
| E | 1 L-R | 78 | 71 |

### 2.2 Patient-specific model reconstruction

The patient-specific geometries were segmented from the MSCT images using Amira-Avizo (Thermo Fisher Scientific, MA) and 3D Slicer (www.slicer.org) softwares (Figure 1). The computational FE meshes were generated using HyperMesh (Altair Engineering, Inc., MI) software (27, 28). The initial state was chosen at mid-systole, where the BAV leaflets are partially open and assumed to be stress-free (29). As shown in Figure 2b, the reconstructed BAV comprised the non-fused leaflet, fused leaflet, the calcification, and the raphe. The complete patient-specific models, as shown in Figure 2c, included the BAV (with raphe and calcification), the aortic root, the ascending aorta, proximal LV, and the myocardium surrounding the aortic root, aortic–mitral curtain, MV and proximal LV/LA endocardial walls.

**Figure 1.**
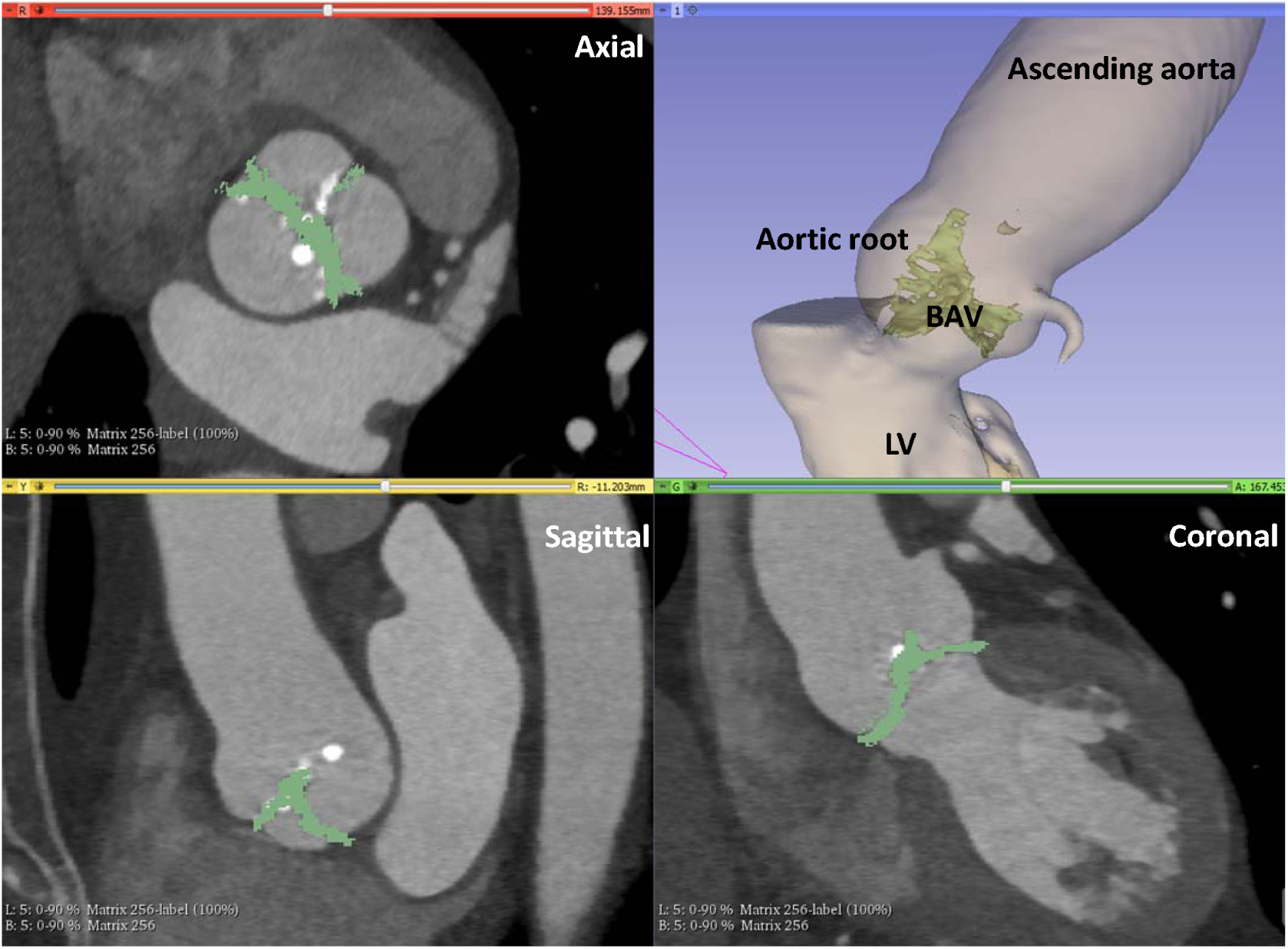
A representative view of the image segmentation in 3D Slicer, the right top panel shows the reconstructed geometry, and the other three panels show the MSCT images in axial, sagittal and coronal views, respectively. The geometry in green represents aortic valve leaflet tissues, the translucent geometry in yellow represents the LV and aorta.

**Figure 2.**
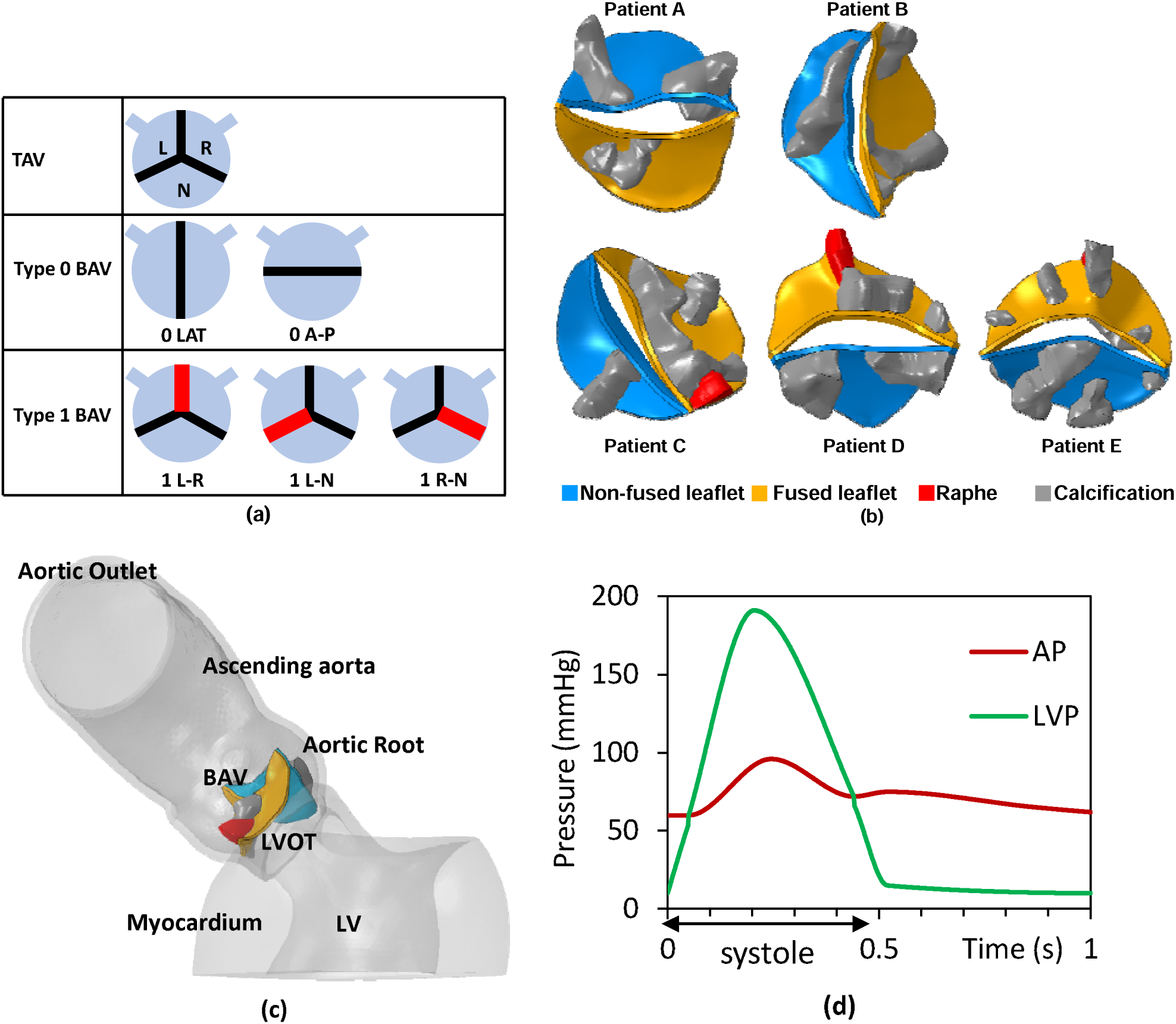
(a) Schematic of normal TAV, type 0 and type 1 BAV. Red line represents the raphe. (b) BAV models that include aortic leaflets, raphe and calcification. (c) Complete patient-specific models (for a representative patient) that include the BAV, aortic root, ascending aorta, proximal left ventricle (LV), and the myocardium. (d) Time-dependent pressure waveforms (for a representative patient) at the LV inlet (LVP) and the aortic outlet (AP).

3D solid elements (eight-node hexahedral C3D8R/C3D8I elements, six-node wedge C3D6 elements, and four-node tetrahedral C3D4 elements) were used to discretize the BAV leaflets, raphe, calcification, aortic root, ascending aorta and myocardium. Three layer of elements were used across the BAV leaflet thickness, with a uniform total thickness of 0.75 mm, which is typical for human AV leaflets (30). Two layers of elements were used across the aorta thickness, with a uniform total thickness of 2□mm. After a mesh convergence study, average mesh sizes for the BAV/raphe, calcification and other components were 0.25 mm, 0.25 mm and 1 mm, respectively. The BAV and calcification shared the same nodes on the tissue-calcification boundary, similarly to the BAV and the aortic root along the leaflet-root attachment curve (ATC). This avoided contact-related and kinematic constraints-related issues during the simulations and ensured full-interface displacement continuity.

### 2.3 FSI modeling of left heart dynamics

A fully coupled FSI numerical approach (31) that combines smoothed particle hydrodynamics (SPH) for the blood flow and nonlinear FE analysis for the heart valve mechanics was implemented into ABAQUS/Explicit (Dassault Systèmes Simulia Corp., Providence, RI, USA) for this study. With the coupling between SPH and FE handled by the node-to-surface contact algorithm, the 3D FSI model was able to simulate the coupled valve nonlinear soft tissue dynamics and the intraventricular hemodynamics in a patient-specific left heart model throughout the whole cardiac cycle. In brief, the continuum medium of blood was discretized as a set of particles distributed over the computational domain without a spatial mesh. The equations of motion and properties of the particles are determined from the continuum equations of fluid dynamics by a kernel interpolation technique (32). In this study, we set a reference density of *ρ* = 1056 *kgm*^−3^ and a viscosity of *μ* = 0.0035 Pa · s for blood properties. SPH particles were initially uniformly distributed in the domain with a spatial resolution of 0.8 mm (33), which led to approximately 1 million one-node (PC3D) elements. The solid domain was reconstructed and discretized using solid elements as described in Section 2.2, and two different constitutive models were used to model the mechanical response of various cardiac tissues. The mechanical response of the ascending aorta, aortic root sinuses, BAV leaflets, raphe and myocardium was modeled with a modified version of the anisotropic hyperelastic Holzapfel-Gasser-Ogden material model (MHGO) (34, 35), in which the cardiac tissues were assumed to be composed of a matrix material with two families of embedded fibers, each consisting of a preferred direction. In addition, the mechanical response of the intervalvular fibrosa was modeled with the isotropic hyperelastic Ogden material model (36). Both constitutive models were implemented into ABAQUS/Explicit with a user sub-routine VUANISOHYPER (37, 38). More details of the modeling of soft tissue materials can be found in the Appendix. The calcification was assumed to be homogeneous, isotropic and linear elastic with a Young’s modulus of 12.6□MPa and a Poisson’s ratio of 0.3 (39).

To simulate the left heart dynamics, time-dependent patient-specific pressure boundary waves (Figure 2d) were applied at the LV inlet and the aortic outlet (31, 40, 41) (Figure 2c), where the mean systolic and diastolic aortic pressures matched the patient data (Table 1). The ascending aorta and myocardium were constrained at their distal ends allowing only rotational degrees of freedom. The initial state was chosen at mid-systole where the leaflets were partially open and assumed stress-free. The patient-specific cardiac cycle duration was calculated using the patient’s heart rate. Two cardiac cycles were simulated and the results from the second cycle were analyzed.

### 2.4 Quantitative analysis of flow field

Three parameters are introduced to analyze the flow field obtained from the FSI simulations quantitively. Firstly, to quantify the flow rotational motion, the vorticity *ω* of the 3D velocity field is calculated using the curl of the velocity field

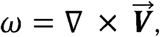

where 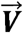 represents the 3D velocity vector field. The magnitude of vorticity represents the local strength of rotational motion, and the direction of vorticity represents the orientation of the local rotation axis. Secondly, the strength of the reverse flow across a cross-sectional plane *S* can be quantified using the flow reversal ratio, defined as

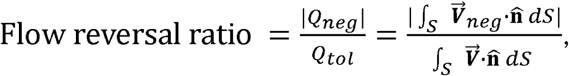

where 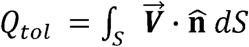 represents the total flow rate, 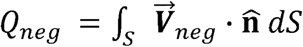 represents the reverse flow rate, with 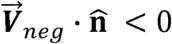, and 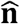 is the normal vector of plane *S*. Lastly, to quantify the helical flow motion in the ascending aorta, the local helicity index (42) is calculated as

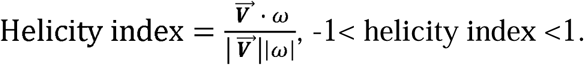

The local helicity index (between -1 and 1) describes the alignment between the velocity direction and the rotation axis orientation. The helicity index is 1 (or -1) when the flow is purely helicoidal where the direction of the velocity vector matches that of the vorticity vector, and the helicity index is 0 when the flow is purely axial (along the centerline axis of aorta) or purely circumferential (within the cross-sectional plane). The positive and negative signs of the helicity index indicate the right-handed and left-handed helical motion, respectively.

### 2.5 Flow metrics of the systolic jet

Three flow metrics were used to characterize the systolic jet. The first one is the peak velocity, which typically occurs at the central core region of the jet. The other two metrics are the normalized flow displacement d’ and the jet angle *θ*, which are defined on the cross-sectional plane *S* immediately downstream of the BAV (Figure 3). The normalized flow displacement d’ = d/R (10) describes the eccentricity of the jet. The distance *d* is between point *C*_*g*_ (geometric center of the plane), and point *C*_*V*_ (location of mean jet velocity through the plane) calculated as

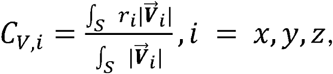

where *r*_*i*_ is the location of the corresponding velocity vector 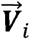. To obtain the normalized flow displacement *d* ‘, the displacement *d* is normalized with the nominal lumen radius R. The normalized flow displacement *d* ’ is between 0 and 1, with a higher value of *d* ’ indicates a more eccentric jet. Lastly, the jet angle *θ* describes the orientation of the jet, which is defined as the angle between the mean jet velocity direction and the normal direction of the plane *S*. A higher value of *θ* indicates more deviation in the jet direction from the centerline axis of the aorta.

**Figure 3.**
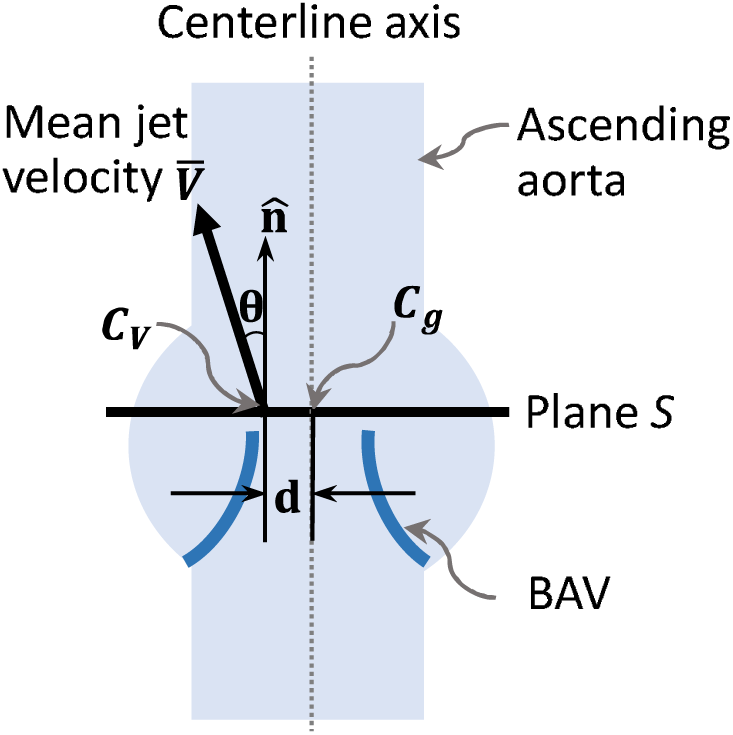
Schematic of the flow displacement *d* and the jet angle *θ*. Vector 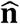 is the normal vector of cross-sectional plane *S*, and vector 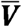 is the mean jet velocity. The flow displacement *d* is the distance between the geometric center of the slice (*C*_*g*_) and the location of mean jet velocity through plane S (*C*_*V*_). The jet angle *θ* is between the normal vector 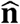 direction and the mean jet velocity 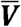 direction. Gray dotted line represents the centerline axis of aorta.

## 3. RESULTS

### 3.1 Model validation

To validate the simulations quantitatively, both the deformed geometries of BAV leaflets and the hemodynamic parameters were compared with the available clinical data. The leaflet location from FSI simulations were extracted at mid-diastole, and compared with the ground truth leaflet geometries segmented directly from the MSCT images. As shown in Figure 4, the deformed BAV geometries obtained from simulations agreed well with the ground-truth geometries, with the mean point-to-mesh error distance less than 1.1 mm for all patients. In addition, as shown in Table 2, the peak systolic velocity obtained from simulations agreed well with the patient’s echo measurement (considered as the ground truth). Moreover, while the clinical data did not report the quantitative value of the AI regurgitant volume, the AI severity predicted from the simulations agreed with that in the clinical report. These validations demonstrated that the coupled SPH-FE FSI models were able to accurately simulate the patient-specific BAV pathological dynamics.

**Figure 4.**
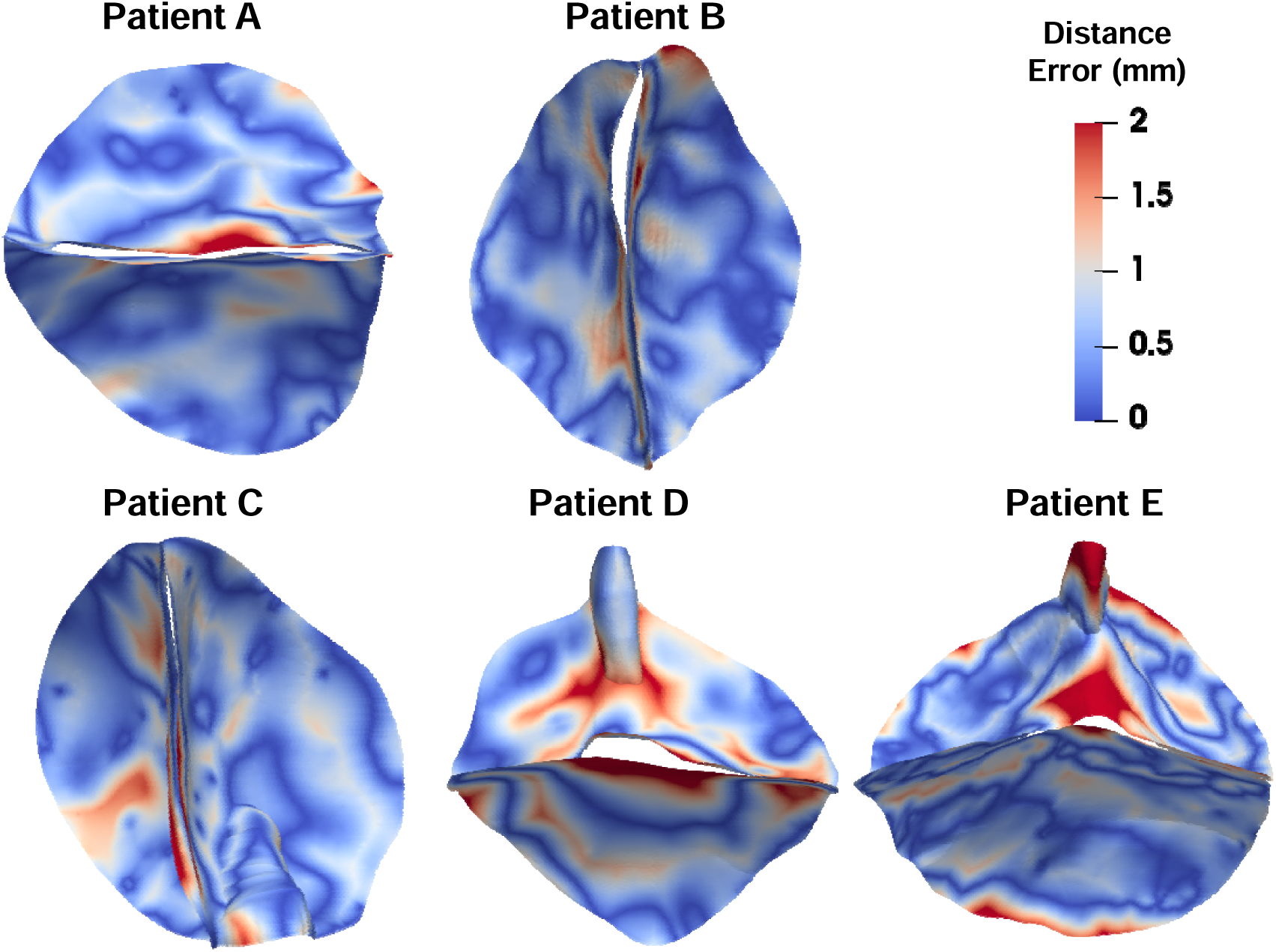
Point-to-mesh distance error between the deformed geometries of the BAV obtained from FSI simulations and the ground truth geometries reconstructed from MSCT images at mid-diastole. Calcifications were not included for comparison. The mean distance errors and the standard deviation (SD) were 0.66 ± 0.63 mm, 0.69 ± 0.64 mm, 0.67 ± 0.59 mm, 1.07 ± 0.92 mm, and 0.64 ± 0.60 mm for patients A, B, C, D, and E, respectively.

**Table 2.** Validation of the peak velocity between the FSI simulations and the clinical data

|  | Patient ID | A | B | C | D | E |
| --- | --- | --- | --- | --- | --- | --- |
| Peak systolic velocity (m/s) | FSI simulation | 5.24 | 5.79 | 3.94 | 4.26 | 3.64 |
|  | Clinical report | 5.20 | 5.25 | 4.00 | 4.73 | 4.20 |

### 3.2 Overall BAV hemodynamics

Figure 5 presents the streamlines across the BAV and within the ascending aorta for the five BAV patients at four time points over the cardiac cycle. During systole, the flow downstream of the BAV can be characterized with two structures: the high velocity systolic jet across the BAV, and the recirculation flow where recirculation vortices locate. At early systole (Figure 5a), the peak velocity of the jet was 1.0-1.5 m/s. The streamlines of the forward flow within the ascending aorta were nearly parallel, and the two counter-rotating recirculation vortices had similar sizes. At peak systole (Figure 5b), the peak velocity of the jet increased significantly to 3.6-5.8 m/s. The systolic jet was typically eccentric, where the center of the jet deviated from the center of the aortic root, and the orientation of the jet deviated from the centerline axis of the ascending aorta. The two recirculation vortices expanded into the ascending aorta and became highly asymmetric. Moreover, the helical flow structure was observed in the ascending aorta. Among the five BAV patients, patient B showed the strongest helical flow structure. At late systole (Figure 5c), the peak velocity of the jet reduced to around 1 m/s, the recirculation vortices shrank back into the sinus, and the helical flow in the ascending aorta became much weaker. During diastole, the flow was in general weak as the BAV closed (Figure 5d). Due to the mild AI, weak retrograde flow was observed across the BAV. The velocity in the ascending aorta was typically less than 0.1 m/s, and the velocity of the regurgitant jet downstream of the BAV was typically less than 0.5 m/s.

**Figure 5:**
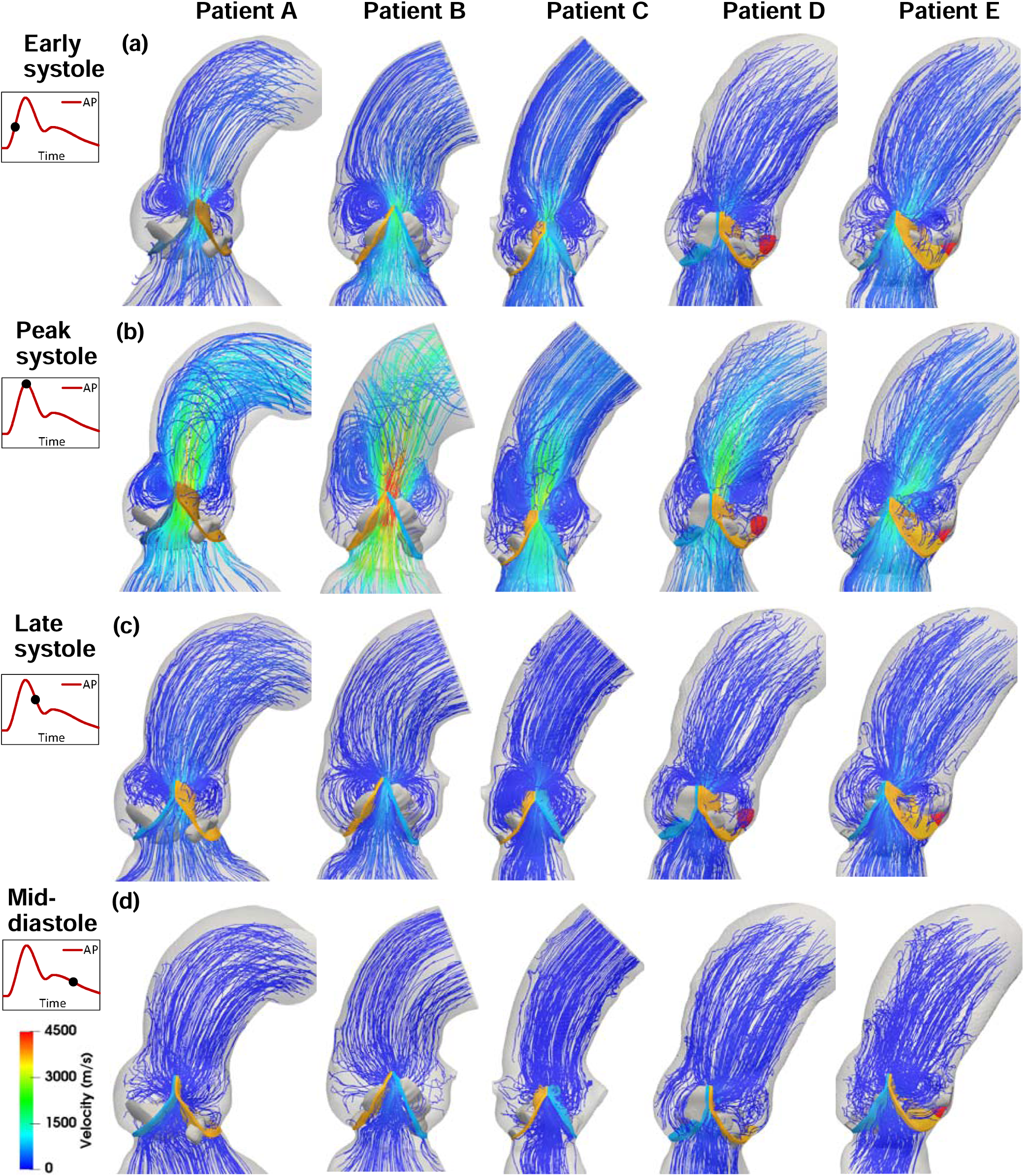
Streamlines of the flow field (side view) obtained from the FSI models for the five BAV patients at four time points: (a) early systole, (b) peak systole, (c) late systole and (d) mid-diastole over the cardiac cycle. The four time points are labeled on the aortic pressure (AP) curve.

To investigate the flow in sinus region, the maximum velocity throughout the entire cardiac cycle was extracted at each location of the flow domain. As shown in Figure 6, outside of the systolic jet region, the maximum velocity in the recirculation flow region was significantly slower (less than 0.3 m/s). The flow on the aortic side the BAV was much slower than that on the ventricular side, and the flow near the ATC at lower sinus was slower than that near the free edges at upper sinus. The potential regions of flow stasis (black colored regions in Figure 6) were observed between the lower sinus and the aortic side of BAV leaflets, where the maximum velocity is less than 0.1 m/s throughout the cardiac cycle.

**Figure 6:**
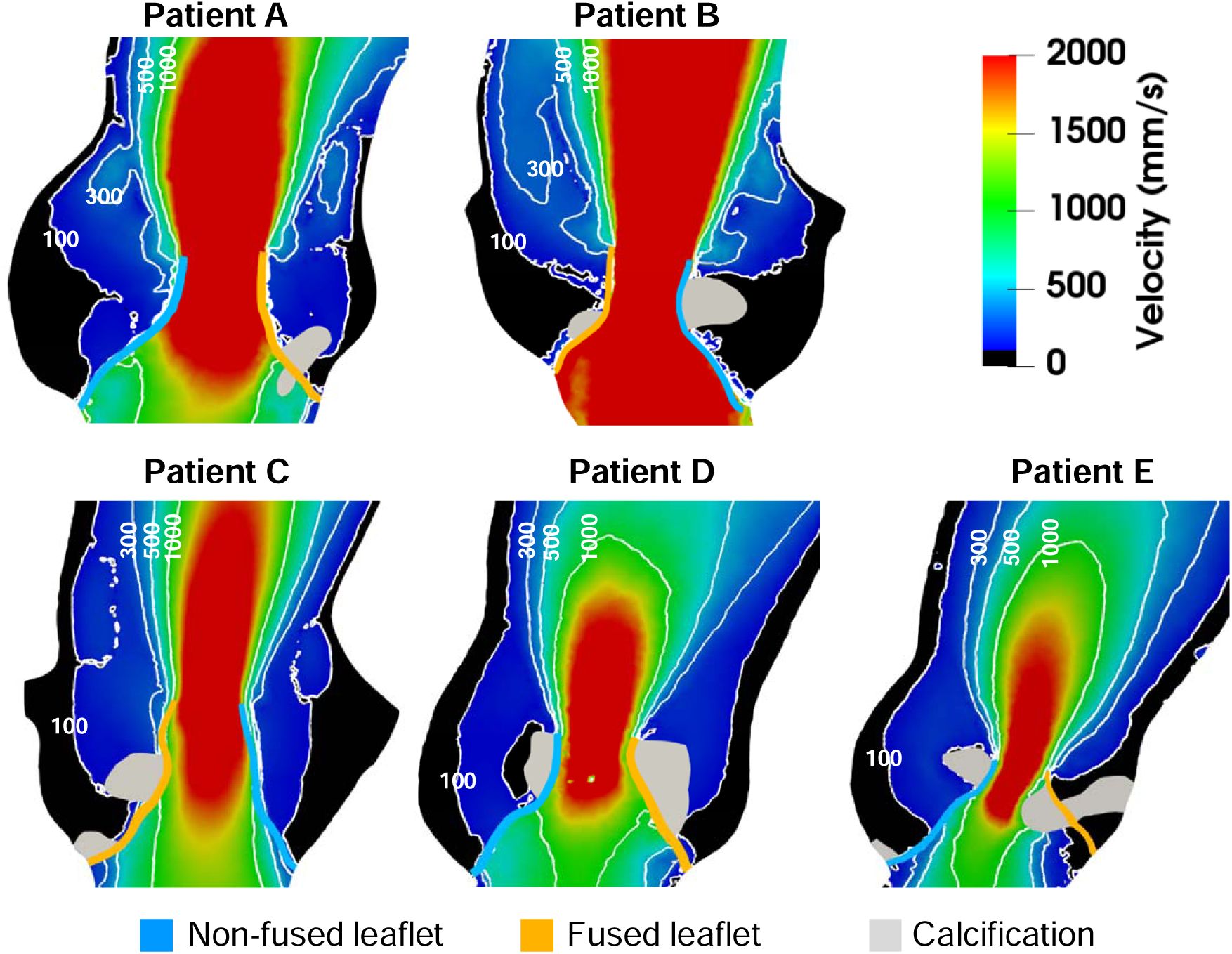
Flow velocity contour plots showing the maximum velocity within the aortic root over the entire cardiac cycle, overlapped with the BAV leaflet geometries at peak systole. The velocity values are in units of mm/s. The regions where the maximum velocity is less than 0.1 m/s through the entire cardiac cycle are colored in black, indicating potential flow stasis region.

### 3.3 Flow at peak systole

For the flow at peak systole, Figure 7 presents the velocity contours and velocity vectors at four different cross sections (sinus, STJ, L1, L2, depicted in Fig 7c) from the aortic root to the ascending aorta. At the sinus level, the peak velocities of the systolic jet were observed in the narrow “fish-mouth” shaped (instead of nearly round for a tricuspid aortic valve) regions, which corresponded to the stenotic opening of BAV. At the STJ level, peak velocity regions started to deviate from the center of aorta, and the velocity direction started to deviate from the normal direction of the cross section. Within the aortic root (sinus and STJ level), forward flow was observed in the center region, and reverse flow was observed near the aortic walls, with the presence of the recirculation vortices (Figure 5b). As the systolic jet exited the aortic root and entered the ascending aorta (L1 and L2 level), the peak velocity started to decrease and the reverse flow gradually weakened. On the other hand, the in-plane secondary flow became more evident. Patient-specific variations were observed among the five patients. Specifically, patient B had the most severe AS with fastest eccentric systolic jet, and the strongest helical flow motion.

**Figure 7:**
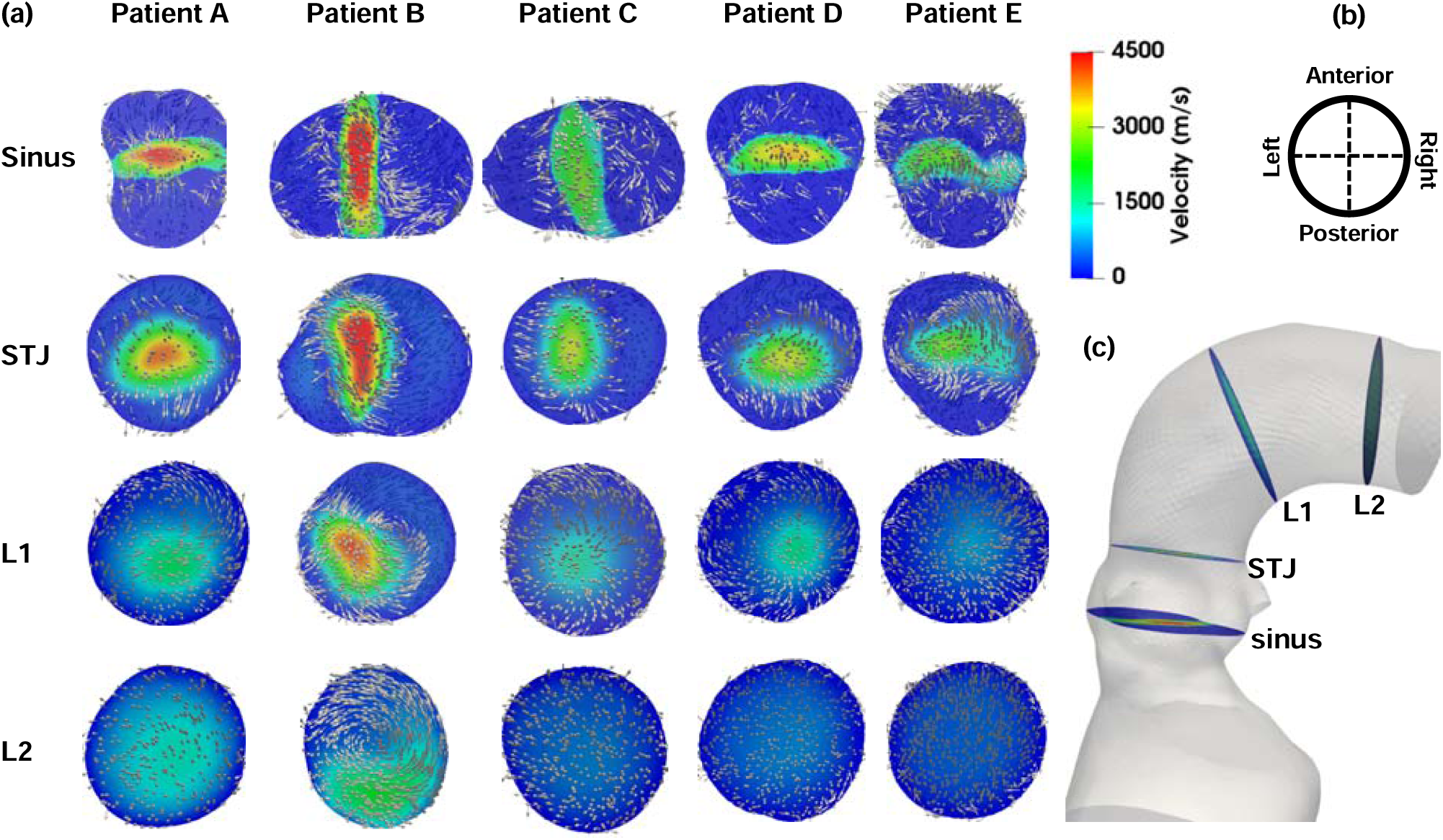
(a). Flow field at four different cross sections from the aortic root to the ascending aorta at peak systole for the five BAV models. Both the velocity contour and the velocity vectors are plotted. (b) The alignment of cross sections. (c) The four cross sections are at the level of the sinuses of Valsalva, the sinotubular junction (STJ), L1, and L2, where L2 is close to the aortic arch, and L1 is roughly in the middle between STJ and L2.

To quantify the flow structure at peak systole, the magnitude of vorticity, flow reversal ratio and the absolute helicity index over four cross sections (sinus, STJ, L1, and L2) were calculated (Table 3). Figure 8 presents the mean ± SD values averaged among five patients at each cross section. In general, the magnitude of the averaged vorticity was the largest (263.1 ± 56.2 s^-1^) at the sinus, and gradually decreased along the aorta (234.0 ± 58.1 s^-1^ at STJ, 119.6 ± 62.2 s^-1^ at L1, and 91.0 ± 42.2 s^-1^ at L2). The flow reversal ratio was 0.013 ± 0.006 at the sinus, increased to the maximum value of 0.074 ± 0.035 at the STJ where the expanded recirculation vortices located (Figure 5b), and then decreased sharply along the ascending aorta (0.017 ± 0.033 at L1, and 0.0006 ± 0.0005 at L2). The helicity index did not vary significantly along the aorta, the mean value increased slightly from the sinus (0.21 ± 0.02) to the STJ (0.22 ± 0.03), and to L1 (0.25 ± 0.05), then slightly decreased downstream (0.22 ± 0.10 at L2).

**Table 3:** The mean vorticity magnitude, flow reversal ratio and the helicity index at different cross sections (sinus, STJ, L1, and L2) for all five BAV patients. The values were averaged over each cross section. For the helicity index, the absolute values were averaged, which represented the overall strength of the helical flow.

|  | Patient ID<br>Location | A | B | C | D | E |
| --- | --- | --- | --- | --- | --- | --- |
| Vorticity<br>($s^{-1}$) | Sinus | 309.62 | 335.02 | 233.73 | 261.06 | 176.11 |
|  | STJ | 311.39 | 290.73 | 202.70 | 208.84 | 156.18 |
|  | L1 | 131.70 | 232.64 | 83.84 | 98.85 | 51.10 |
|  | L2 | 129.82 | 152.99 | 60.48 | 64.42 | 47.30 |
| Flow<br>reversal<br>ratio | Sinus | 0.01338 | 0.00979 | 0.02352 | 0.01346 | 0.00578 |
|  | STJ | 0.06893 | 0.13045 | 0.09224 | 0.04310 | 0.03522 |
|  | L1 | 0.00043 | 0.08545 | 0.00057 | 0.00156 | 0.00162 |
|  | L2 | 0.00009 | 0.00002 | 0.00067 | 0.00070 | 0.00129 |
| Helicity<br>index | Sinus | 0.2202 | 0.2262 | 0.2195 | 0.1814 | 0.2055 |
|  | STJ | 0.2081 | 0.2079 | 0.1990 | 0.1937 | 0.2740 |
|  | L1 | 0.3382 | 0.2492 | 0.1964 | 0.2575 | 0.1960 |
|  | L2 | 0.1853 | 0.4171 | 0.1799 | 0.1685 | 0.1640 |

**Figure 8:**
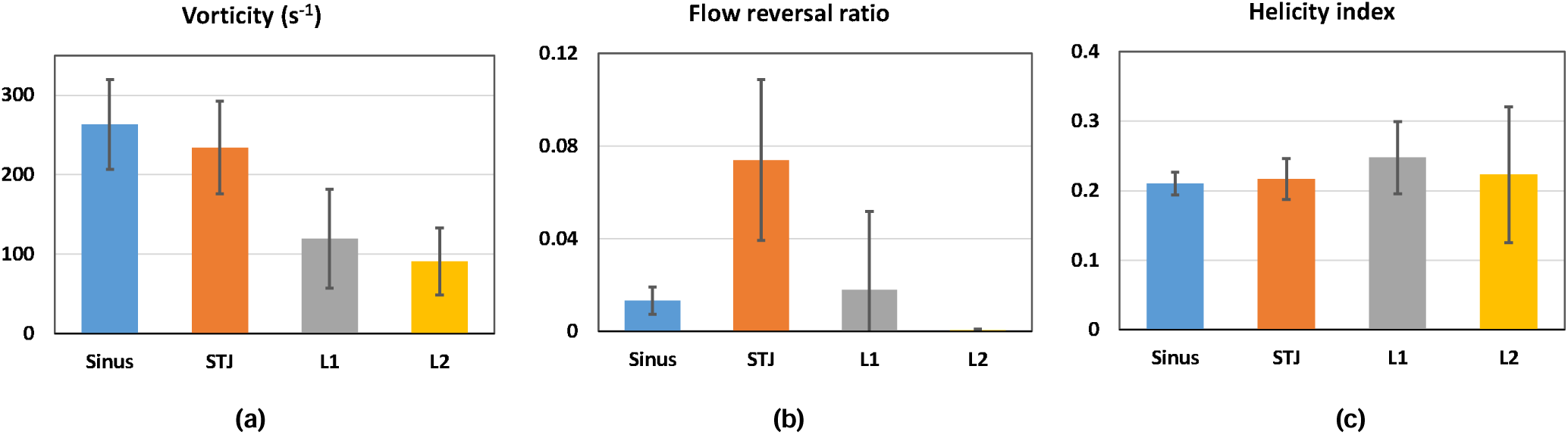
The mean ± SD values of the (a) vorticity magnitude, (b) flow reversal ratio, and (c) absolute helicity index on the four cross sections at the sinus, STJ, L1 and L2. The values were averaged among the five BAV patients.

To characterize the systolic jet, the peak systolic velocity, the normalized flow displacement d’, the jet angle _θ_ were quantified for each patient (Table 4) as described in Section 2.5. Among all five patients, patient B had the fastest systolic jet with high eccentricity.

**Table 4:**
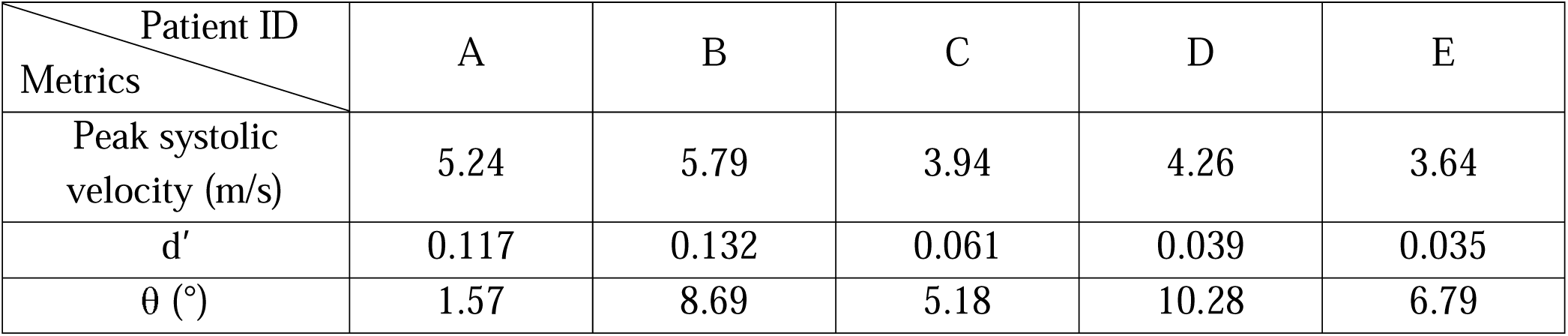
The peak systolic velocity, the normalized flow displacement d’, the jet angle θ for all five BAV patients.

## 4. DISCUSSION

The main contribution of this study is a comprehensive analysis of BAV hemodynamics in rigorously developed patient-specific cardiac computational models. Specifically, this work presents an engineering study that

1. Integrated cardiac tissue mechanics and blood flow modeling using a fully coupled FSI framework which allowed for an accurate assessment and validation of patient-specific BAV hemodynamics.
2. Investigated the hemodynamics of BAV over the entire cardiac cycle using patient-specific models, which included the BAV leaflets (with raphe and calcifications), aortic root, ascending aorta, part of the LV and the surrounding myocardium.
3. Quantified the flow patterns at peak systole using the vorticity, flow reversal ratio and helicity index at different locations from the aortic root to the ascending aorta.
4. Investigated the relation between multiple jet metrics (peak velocity, normalized flow displacement, jet angle) and the flow patterns within the ascending aorta.

### 4.1 Patient-specific analysis of BAV hemodynamics using FSI modeling

#### 4.1.1 BAV hemodynamics throughout the cardiac cycle

During systole, the flow can be characterized as a jet flow through the aortic valve into a confined geometry of the aorta (Figure 9a). The peak velocity at the centerline of the jet remains nearly uniform within the potential core region immediately downstream the aortic valve. As the jet gradually expands, the potential core shrinks, and the shear layer grows within which the entrainment of the quiescent fluid occurs. Within a confined geometry of the aorta, this entrainment drives the formation of recirculation vortices, which circles from the belly region towards the free edge of the BAV leaflets, and turns around along the sinus after the recirculation flow hits the aortic wall.

**Figure 9:**
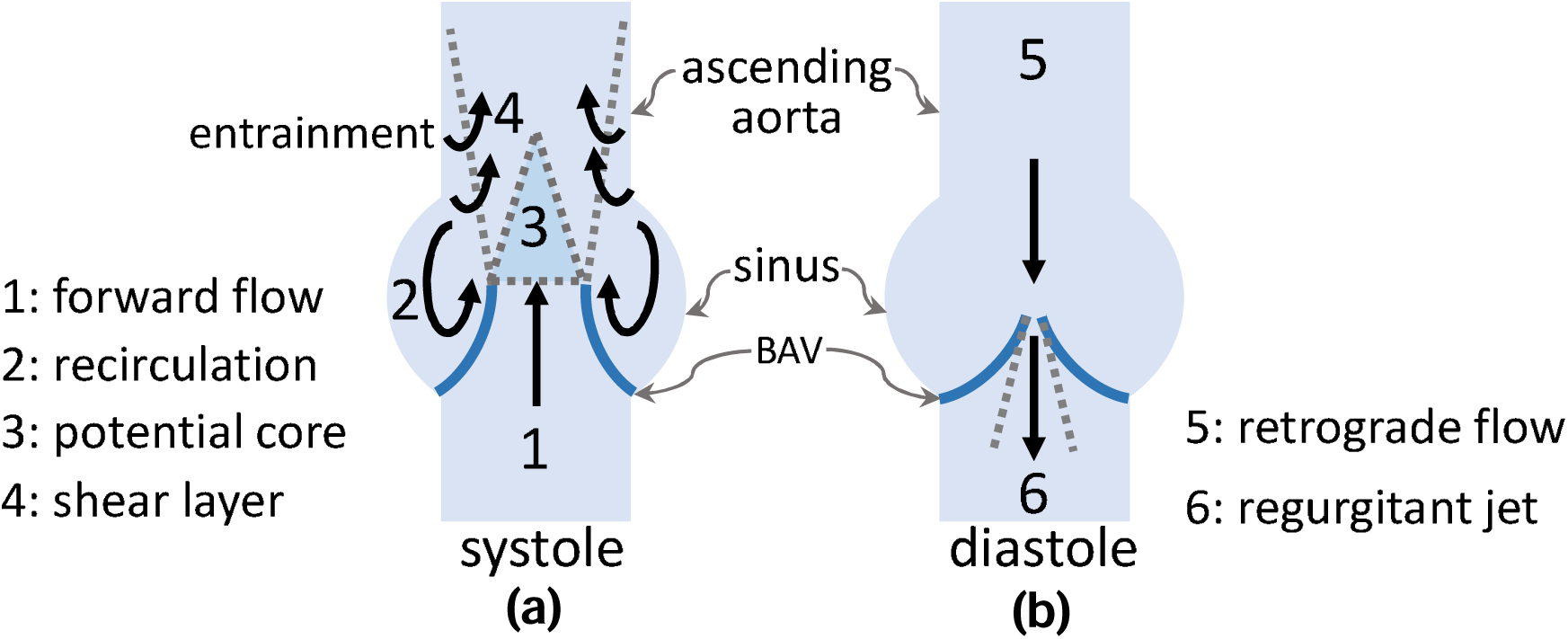
Schematic showing the overall flow structure at (a) systole and (b) diastole. The schematic at diastole corresponds to the BAV patients with mild AI.

During diastole, the flow is in general very weak as the BAV closes. No coherent flow structures are observed as the flow momentum gradually dissipates, which is consistent with previous *in vitro* observations (43). While the mild AI leads to converging retrograde flow towards the BAV and regurgitant jet downstream of the BAV (Figure 9b), the regurgitant jet is much weaker than the systolic jet, and the entrainment within the sinus is too weak to form any coherent recirculation vortices.

#### 4.1.2 Flow in the sinus

The flow within the sinus is slow throughout the cardiac cycle. In the upper sinus, two recirculation vortices typically form during systole, corresponding to the two BAV leaflets. Driven by the entrainment of the systolic jet, the two vortices always rotate in the opposite direction, and these directions do not change over the cardiac cycle. This is consistent with experimental studies (43, 44) which found that the shear stress on the leaflet aortic side was unidirectional rather than bidirectional. In the lower sinus, the flow is significantly slower and nearly stagnant over the entire cardiac cycle (Figure 6). This indicates that the calcium build-up is more likely to occur near the ATC at lower sinus than near the free edges at upper sinus.

#### 4.1.3 Flow at peak systole

The flow structures at peak systole are quantified using the vorticity, flow reversal ratio and the helicity index. The magnitude of the vorticity describes the rotation strength of the flow. Since the rotational motion of the flow is mostly induced by the high velocity systolic jet passing through the BAV leaflets, the vorticity is the largest in the sinus, and gradually decreases within the ascending aorta as the rotational momentum of the flow dissipates (Figure 8a). The flow reversal ratio describes the strength of reverse flow, which is mostly associated with the recirculation vortices. As the result, the reversal flow ratio is the largest at the STJ, and decreases to nearly zero downstream of the ascending aorta (Figure 8b). The helicity index describes the relative strength of the helical motion. Unlike the rotational and reversal motion, the helical motion of the blood flow could be induced by the torsion in the ascending aorta (14). Therefore, the helicity index does not vary much within the ascending aorta (Figure 8c).

The flow complexity is found to be associated with the velocity and eccentricity of the systolic jet. At early and late systole, slower jet leads to weaker entrainment, and the entire flow field is overall well regulated (Figure 5a, 5c). At peak systole, the flow shows richer dynamics as the systolic jet reaches maximum velocity and is usually eccentric (Figure 5b). The recirculation vortices expand due to the stronger entrainment, and the two vortices become asymmetric due to the combined effect of the eccentric jet and the asymmetric leaflet/sinus geometries. Furthermore, helical motion was also observed in the ascending aorta. A more detailed quantitative analysis of correlations between the flow patterns and jet metrics is discussed below in Section 4.2.

#### 4.1.4 Patient-specific BAV hemodynamics

Our results suggest that patient-specific FSI modeling is important for comprehensive analysis of BAV hemodynamics. Previous fluid dynamics analyses that ignored the valves (11, 12) only investigated the flow at peak systole, and were unable to capture the dynamic motion of the jet and recirculation vortices in the sinus. FSI studies that used 2D models (23) were unable to predict the helical flow pattern, which is characteristic of BAV hemodynamics. Furthermore, the studies using idealized geometries were unable to capture accurate patient-specific variations. For example, Marom et al. (45) reported that the size of recirculation vortex was simply associated with the leaflet/sinus size, while we found that the jet eccentricity affected the vortex size as well. In addition, Liu et al. (14) suggested that the torsion of aorta was an important factor that induced the helical flow motion, therefore simulations with an idealized aorta with only in-plane curvature (22) might not be able to accurately predict helical flow motion and secondary flow in cross sections.

### 4.2 Relation between jet metrics and flow patterns in the ascending aorta

To investigate the effects of the systolic jet on the overall flow patterns within the ascending aorta, the parameters that quantified the flow structures (described in Section 2.4) are correlated with different flow metrics of the systolic jet (described in Section 2.5) using the linear regression analysis (Figure 10). The parameters that quantified the flow structures are the mean values of vorticity, helicity index and the flow reversal ratio over the ascending aorta (from STJ to L2 location), while the jet flow metrics include the peak velocity, the normalized flow displacement d’, and the jet angle _θ_. The peak velocity of the systolic jet showed good correlation with the vorticity and helicity index, and an acceptable correlation with the flow reversal ratio (Figure 10a). This suggests that the aortic stenosis could enhance the flow complexity and the helical flow structure in the ascending aorta, which is consistent with previous numerical studies (19) and *in vivo* patient data analysis (46). The normalized flow displacement d’ shows good correlation with the vorticity, helicity index and the flow reversal ratio (Figure 10b), but the jet angle _θ_ does not correlate well with any of these parameters (Figure 10c). These findings are consistent with the previous study of Sigovan et al. (10) which found the normalized flow displacement better characterized the degree of eccentric flow, and the study of Mahadevia et al. (6) who observed that the normalized flow displacement was more sensitive than the jet angle to differences in BAV aortopathy phenotype. Considering that the measurement of jet metrics is more feasible in the clinical practice than the detailed analysis of flow field, our findings suggest that the peak systolic velocity and the normalized flow displacement could be useful noninvasive metrics for identifying and evaluating the abnormal flow structures in the ascending aorta.

**Figure 10:**
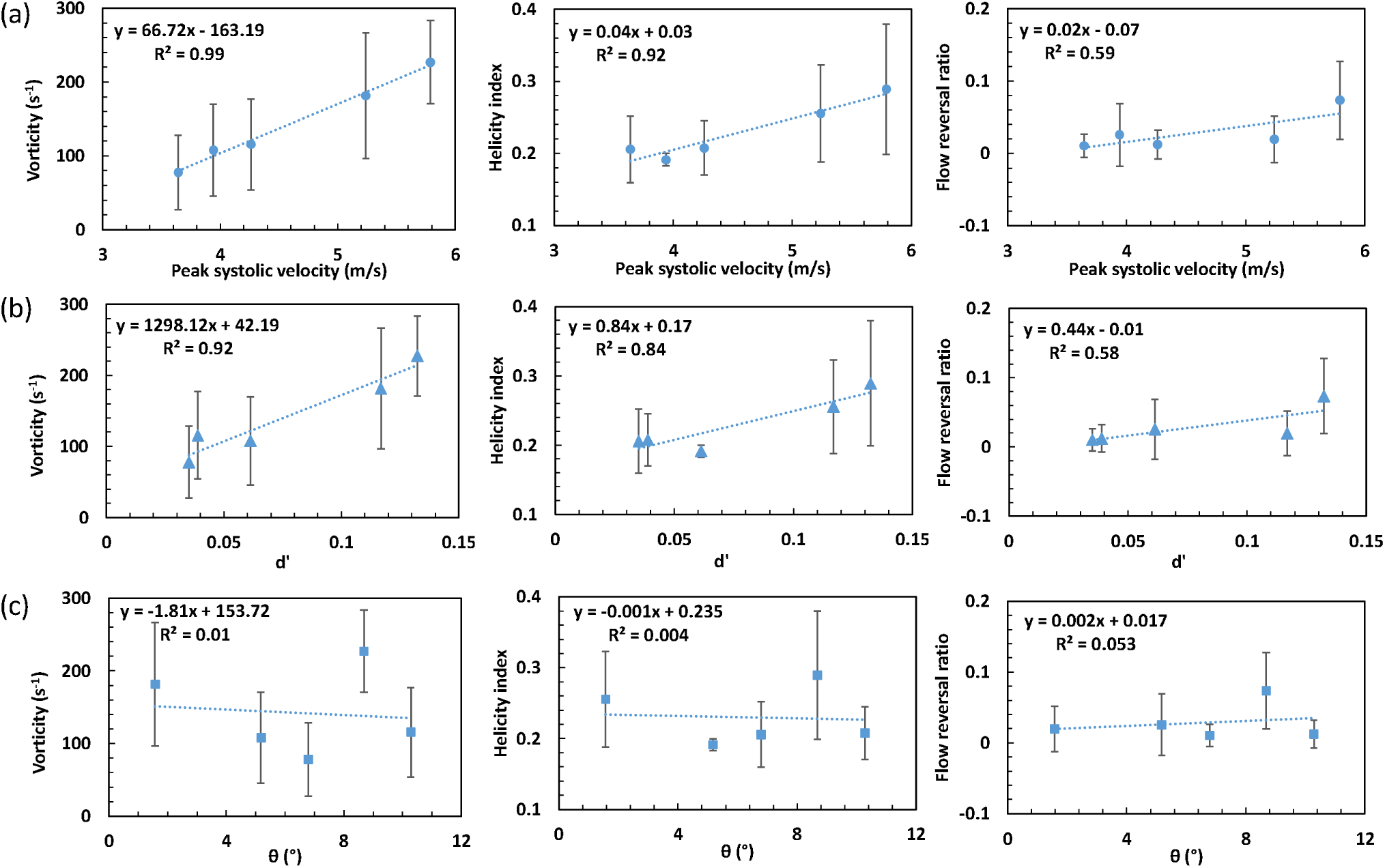
Linear regression analysis between the jet flow metrics (peak systolic velocity, the normalized flow displacement d’, the jet angle _θ_) and the vorticity, helicity index, flow reversal ratio over the ascending aorta. The five data points in each correlation represents the values of the five BAV patients. The mean values of vorticity, helicity index and flow reversal ration of each patient were averaged over three cross sections (STJ, L1, and L2).

### 4.3 Limitations

There are several limitations in this study. Firstly, while the BAV leaflets were deformable, the aorta was essentially stationary. More accurate simulations will consider the pre-stress and the aorta deformation. Secondly, due to the inherent limitation of Abaqus SPH formulation, small-scale flow and turbulence features may not be accurately resolved close to the aortic wall. However, as discussed in previous numerical studies (13, 19, 45), simulations without a turbulent model were able to capture the hemodynamic phenomena of interest in the bulk flow, and the simulation results in this study have been validated with available *in vivo* data. Finally, a limited number of type 0 (n = 2) and type 1 (n=3) BAV patient data were collected. Therefore, it is unfeasible to investigate the effect of BAV phenotypes on BAV hemodynamics. Future studies would benefit from a larger number of patients with the inclusion of different BAV phenotypes.

## 5. CONCLUSIONS

We have performed patient-specific FSI analysis of BAV hemodynamics. The systolic flow can be characterized by a systolic jet and two counter-rotating recirculation vortices, the diastolic flow was weak without coherent flow structures, and the sinus flow was slow throughout the entire cardiac cycle, especially in the lower sinus. The flow at peak systole showed the richest dynamics, with eccentric jet, asymmetric recirculation vortices, and varying helical flow pattern in the ascending aorta. The flow structure at peak systole was quantified using the vorticity, flow reversal ratio and helicity index at multiple locations. While the vorticity and flow reversal ratio decreased significantly from the aortic root towards the ascending aorta, the helicity index did not vary much and typically reached maximum within the ascending aorta. The complex flow structures were found to be associated well with the velocity and the eccentricity of the systolic jet. In specific, both the peak velocity and the normalized flow displacement (rather than the jet angle) of the systolic jet showed a strong correlation with the vorticity and helicity index of the flow downstream in the ascending aorta, which suggests that these jet metrics can be used for noninvasive evaluation of BAV flow structures.

## Data Availability

All data produced in the present work are contained in the manuscript.

## ACKNOWLEDGEMENTS

This study was supported in part by the NIH R01HL104036, and the Marvin H. and Nita S. Floyd Research Fund. Dr. Tongran Qin was in part supported by the American Heart Association (AHA) Postdoctoral Fellowship 19POST34450161. Dr. Andrés Caballero was in part supported by a Fulbright-Colciencias Fellowship. Dr. Wenbin Mao was in part supported by the AHA 19CDA34660003 grant.

## Appendix

### Modeling of mechanical material properties for Cardiac tissues

The mechanical response of the ascending aorta, aortic root sinuses, BAV leaflets, raphe and myocardium was modeled with a modified version of the anisotropic hyperplastic Holzapfel-Gasser-Ogden material model (MHGO) (34, 35), in which the cardiac tissues were assumed to be composed of a matrix material with two families of embedded fibers, each consisting of a preferred direction. The strain energy function, *W*, can be expressed as

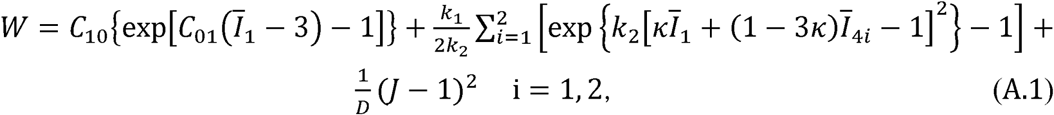

where *c*_1 0_,*c*_0 1_ are the matrix parameters, *k*_1_,*k*_2_ are the material parameters, *D* is a material constant which enforces incompressibility, *Ī*_1_ and *Ī*_4*i*_ are the strain invariants (47-49), which describe the matrix material and the fiber family properties, respectively, *K* is a dispersion parameter which determines the level of dispersion in the fiber orientations, and *J* is the determinant of the deformation gradient. The mean local fiber directions are assumed symmetric with respect to the circumferential axis of the local coordinate system. Local coordinate systems were defined for each leaflet, and the local fiber orientations were defined through 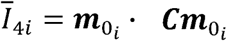, where ***C*** is the right Cauchy Green tensor, with 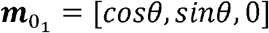, and 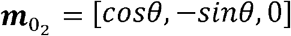 and *θ* defines the angle between one of the mean local fiber direction and the circumferential direction of the local coordinate system.

In addition, the mechanical response of the intervalvular fibrosa was modeled with the isotropic hyperelastic Ogden material model (36).

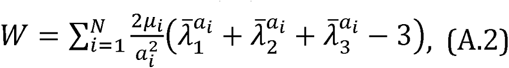

where μ_*i*_ and *a*_*i*_ are material constants, and 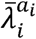 are the modified principal stretches. The anisotropic hyperelastic MHGO model and the isotropic hyperelastic Ogden model were implemented into ABAQUS/Explicit 2016 (Dassault Systèmes Simulia Corp., Providence, RI, USA) with a user sub-routine VUANISOHYPER (37, 38), where the model parameters (Table A.1) were obtained from the in-house multiprotocol biaxial and uniaxial testing data of human cardiac tissues in our lab. The calcification was assumed to be homogeneous, isotropic and linear elastic, with a Young’s modulus of 12.6□MPa and a Poisson’s ratio of 0.3 (39). SPH particles were given Newtonian blood properties with a density of *ρ* = 1056 kg/*m*^3^ and a dynamic viscosity of *µ* = 0.0035 Pa · s.

For a normal TAV patient, the collagen fibers on the leaflets are typically orientated circumferentially from commissure to commissure, and nearly parallel to the free edge (50-52). However, for a fused leaflet with raphe, the fibers in the raphe region are typically disoriented (53, 54). It has been reported that the orientation of the fibers can be almost 45° (instead of parallel) to the free edge, and in the raphe, the fibers from the opposite side can merge at a 90° angle (53). Therefore, the local coordinate system was defined accordingly (25): for the leaflet without raphe, the circumferential axis of the local coordinate system was set to be nearly parallel to the free edge; while for the leaflet with raphe, the circumferential axis was set to be approximately +45° and -45° to the free edge on both sides of the raphe, forming a 90° angle at the raphe.

**Table A.1.** The cardiac tissues material parameters used in the MHGO material model and the Ogden model.

| **MHGO model** | $C_{10}$ (kPa) | $C_{01}$ | $k_1$ (kPa) | $k_2$ | $\theta$ ($^\circ$) | $\kappa$ | $D$ (kPa$^{-1}$) |
| --- | --- | --- | --- | --- | --- | --- | --- |
| BAV leaflets and raphe | 0.017 | 147.262 | 39704.1 | 2352.96 | 0 | 0.317 | 5.0e-4 |
| Sinuses | 1.755 | 13.707 | 10.550 | 80.379 | 20.06 | 0 | 5.0e-4 |
| Aorta | 4.175 | 3.464 | 3.771 | 15.927 | 70.95 | 0.086 | 5.0e-4 |
| Myocardium | 0.0374 | 15.387 | 6.079 | 98.366 | 6.78 | 0.144 | 5.0e-4 |
| **Ogden model** | $\mu_1$ (MPa) | $a_1$ | $\mu_2$ (MPa) | $a_2$ | $\mu_3$ (MPa) | $a_3$ | |
| Intervalvular fibrosa | 1.505 | 21.400 | 11.207 | 21.400 | 1.441 | 21.400 |  |

